# Unexpectedly higher levels of naturally occurring anti-Orthopoxvirus neutralizing antibodies are observed among gay men than general adult population

**DOI:** 10.1101/2022.12.20.22283644

**Authors:** Yanmeng Feng, Yifan Zhang, Shengya Liu, Cuiyuan Guo, Wanhai Wang, Wenhong Zhang, Heng Tang, Yanmin Wan

**Affiliations:** Hubei Provincial Center for Disease Control and Prevention, Wuhan 430065, China; Department of Infectious Disease of Huashan Hospital, National Medical Center for Infectious Diseases and Shanghai Key Laboratory of Infectious Diseases and Biosafety Emergency Response, Fudan University, Shanghai 200040, China; Clinical Laboratory, The First Affiliated Hospital of Zhengzhou University, Key Laboratory of Laboratory Medicine of Henan Province, Zhengzhou 450052, China; Shenzhen International Travel Health Care Center (Shenzhen Customs District Port Outpatient Clinics), Shenzhen Customs District, Shenzhen 518033, China; State Key Laboratory of Genetic Engineering, School of Life Science, Fudan University, Shanghai 200438, China; Department of radiology, Shanghai Public Health Clinical Center, Shanghai 201508, China

## Abstract

The confirmed cases in the current outbreak of Monkeypox are predominantly identified in the networks of men who have sex with men (MSM). It is suggested that special behavioral characteristics might make the virus spread more easily in this population, but the biological factors affecting the spread of this outbreak have not been fully clarified. In this study, we measured the anti-Monkeypox and anti-Vaccinia antibodies in an MSM cohort (comprising 326 individuals) and a general population cohort (comprising 295 individuals) and compared the antibody responses between the two cohorts. Meanwhile, we also compared the antibody responses between individuals born before and in/after 1981, when the smallpox vaccination was ceased in China. Our data showed that binding antibodies against Monkeypox H3L, A29L, A35R proteins and Vaccinia whole-virus lysate could be detected in individuals born both before and in/after 1981, of which the anti-Vaccinia binding antibody levels were found to be significantly higher among the individuals born before 1981 in the general population cohort. Moreover, we unexpectedly found that the levels of binding antibody responses against Monkeypox proteins were significantly lower among individuals of the MSM cohort born in/after 1981, but the anti-Vaccinia neutralizing antibody levels were significantly higher among these individuals compared to those age-matched participants of the general population cohort. Additionally, we demonstrated that the positive and negative rates of anti-Monkeypox antibody responses were associated with the anti-Vaccinia antibody responses among individuals born before 1981 in the general population cohort, but no significant association was observed among individuals born in/after 1981 in both cohorts. Further studies are warranted to clarify the impact of the naturally occurring anti-Orthopoxvirus antibodies on the transmission of Monkeypox, especially among gay men who have not been vaccinated against smallpox.

## Introduction

Monkeypox virus (MPXV) is a DNA virus in the *Orthopoxvirus* genus, which was first discovered in 1959 when cynomolgus macaques were shipped from Singapore to a Denmark research facility [1, 2]. Despite being named after the first described host, monkeys and humans are accidental hosts for MPXV, its natural hosts are more likely to be rodents [3, 4]. MPXV comprises two distinct lineages, the Western Africa strain and the Congo Basin strain. Sporadic human infections by both strains have been reported previously, of which the Congo Basin strain causes more severe disease [5, 6] and accounts for more cases [7].

Compared to previous observations, which showed that the ability of MPXV to transmit among humans was weak [8] and the major clinical manifestations included fever, skin rash and swollen lymph nodes [7], the emerging outbreak is unusual in the following three aspects. First, the current outbreak spreads faster than ever before. Since the first case was reported on May 7, 2022, over 82000 confirmed cases have been reported (including 65 deaths) by December 14, 2022, spanning 110 nations and regions.

Second, it has been shown in the current outbreak that MPXV can present symptoms typical for sex transmitted infections, such as urethritis, rectal pain and urinary retention [9]. Third, there was a link between MPXV transmission and sexual contact among men who have sex with men [10-13]. Moreover, it is also surprising that the virus causing the current outbreak belongs to the Western African lineage [14], which historically demonstrated low outbreak-causing potential [6, 15].

Epidemiological investigations estimated that the basic reproduction number (R_0_) for MPXV-2022 may be substantially above 1 [16, 17], but the underlying determinants of the outbreak remain elusive [18]. Factors, such as the waning immunity to smallpox [7], genetic evolution of virus [14, 19] and super spreader events [16] have been proposed to contribute to the resurgence and the increased human-to-human transmission of MPXV. Although a modeling study showed that smallpox vaccination conferred protection against all Orthopoxviruses [20], it is not the only source of anti-Orthopoxvirus immunity. Two previous studies showed that anti-Orthopoxvirus antibodies could also be detected in significant percentages of individuals either without previous exposures [21] or with low exposure chances [22]. A more recent study suggested that neutralizing antibodies against MPXV could be detected in individuals without previous vaccination or infection [23]. The preexisting antibodies may profoundly impact the transmission of MPXV, however the current-day prevalence of antibodies against any Orthopoxvirus is unknown. To fill the knowledge gap, in this study, we detected and compared the anti-MPXV and anti-Vaccinia in an MSM cohort and a general population cohort.

## Materials and methods

### Study Populations and Sample Collection

In this study, we enrolled a cohort of gay men (n=326) who were under routine follow-up at Hubei Provincial Center for Disease Control and Prevention, China and a cohort of general adults (n=295). Individuals with chronic diseases (tumor, hypertension, coronary heart disease, autoimmune diseases and diabetes) and active infections (Influenza, SARS-CoV-2, HIV, HBV, HCV, tuberculosis, etc.) were excluded. Written informed consents were obtained from all participants, and the study was supervised by the Research Ethics Committee of Hubei CDC. The demographical characteristics of the enrollments were depicted in Table 1.

**Table 1.** Demographical characteristics of enrolled participants

|  | Number | Age<br>(Median, 25%-75% percentile) |
| --- | --- | --- |
| **General_Man** |  |  |
| ≤41years | 46 | 33.00 (28.00-37.25) |
| >41years | 72 | 53.00 (48.00-60.75) |
| **General_Woman** |  |  |
| ≤41years | 97 | 31.00 (28.00-36.00) |
| >41years | 80 | 49.00 (45.00-55.50) |
| **MSM** |  |  |
| ≤41years | 321 | 26.00 (23.00-29.00) |
| >41years | 5 | 48.00 (43.50-52.00) |

**Note:** MSM, men who have sex with men.

Peripheral blood was collected by venipuncture into ethylenediaminetetraacetic acid (EDTA) tubes by medical personnel. After centrifugation, plasma was transferred into cryotubes and stored at −80°C until tested.

### Detection of binding IgG antibodies against MPXV H3L, A29L and A35R proteins

MPXV H3L, A29L and A35R proteins were genetically similar with Vaccinia virus H3, A27 and A33 proteins, respectively [24]. As Vaccinia virus proteins A27, H3 and A33 are known to elicit neutralizing antibodies [25] and immunization with A27, A33 [26] or H3 [27] protected against challenge with virulent virus in mice or macaques, we speculated that antibodies against MPXV H3L, A29L and A35R might be relevant to protection either. In-house enzyme-linked immunosorbent assays (ELISA) were developed to measure binding antibodies against MPXV H3L, A29L and A35R proteins. Briefly, high-binding 96-well EIA plates (Cat# 9018, Corning, USA) were coated with purified MPXV A35 (Cat# 40886-V08H, Sino Biological, China), A29 (Cat# 40891-V08E, Sino Biological, China) or H3 (Cat# 40893-V08H1, Sino Biological, China) proteins at a final concentration of 1μg/ml in carbonate/bi-carbonate coating buffer (30mM NaHCO_3_,10mM Na_2_CO_3_, pH 9.6). Subsequently, the plates were blocked with 1×PBS containing 5% skimmed milk for 1 hour at 37°C. Then, 50μl of 1:50 diluted human plasma was added to each well. After 1-hour incubation at 37°C, the plates were washed with 1×PBS containing 0.05% Tween20 for 5 times and incubated with 0.5M NaSCN for 15 minutes at room temperature. After incubation, the plates were immediately washed for 2 times. Next, 50μl of an HRP labeled goat anti-human IgG antibody (Cat# ab6759, Abcam, UK) diluted in 1×PBS containing 5% skimmed milk were added to each well and incubated for 1 hour at 37°C. After another round of wash, 50μl of TMB substrate reagent (Cat# MG882, MESGEN, China) was added to each well. 25 minutes later, the color development was stopped by adding 50μl of 1M H_2_SO_4_ to each well and the values of optical density at 450nm and 630nm were measured using a microplate reader (Cat# 800TS, Biotek, USA). The cut-off value was defined as 3-fold of the average OD_450nm-630nm_ of wells coated with coating buffer.

### Detection of binding IgG antibodies against Vaccinia whole-virus lysate

Vaccinia Tiantan virus suspended in 1×PBS were boiled for 10 minutes and the concentration of total protein was quantified using a bicinchoninic acid assay (BCA) (Cat# E112-01, Vazyme, China) according to the manufacturers’ instruction. The experiment procedure was the same with the in-house ELISA assays described above, except that the coating antigen was replaced with Vaccinia whole-virus lysate, which was used to coat the EIA plates at a final concentration of 20μg/ml.

### Plaque Reduction Neutralization Test (PRNT)

The method of PRNT was established according to published literatures [28, 29] with minor modifications. Briefly, test sera were 1:15 diluted in Dulbecco’s modified Eagle’s medium (DMEM) supplemented with 1% FBS and mixed with an equal volume of Vaccinia Tiantan virus suspension (20 pfu/100 μL). The mixtures were incubated at 37°C for 1 hour. After incubation, the mixtures were transferred onto confluent Vero cell monolayers in 24-well cell culture plates. The plates were incubated at 37°C for 2 hours in a humiliated incubator with 5% CO_2_. Next, 200μl of overlay-medium (DMEM containing 1% FBS and 1.25% methyl cellulose) was added to each well after removing the supernatant. The plates were incubated for 4 days at 37°C in the incubator. At the end of incubation, the plates were stained with crystal violet and the number of plaques were manually counted. Fetal bovine serum was used as the negative control. All samples were detected in duplicated wells. The viral inhibition ratio was calculated using the following formula: (The average plaque number of the negative control wells − the average plaque number of the sample wells) / The average plaque number of the negative control wells.

## Statistical analysis

Statistical analyses were conducted using Graphpad Prism 9 (GraphPad Software, USA). Normality tests were performed before all downstream statistical analyses except the Chi square test. Comparisons between two groups were performed by the method of non-parametric *t*-test. Differences among multiple groups were compared by the method of one-way ANOVA. The contingency analysis was done using the method of Chi-square test. Correlation analysis was conducted by the method of Pearson correlation. P ≤ 0.05 was considered as statistically significant.

## Results

### Levels of binding antibody responses against MPXV H3L, A29L and A35R proteins were significantly lower in the MSM cohort than in the general population cohort

To reveal the prevalence of preexisting antibody responses against MPXV in an MSM cohort and a general population cohort, we first measured the binding IgG responses against MPXV H3L, A29L, A35R proteins and Vaccinia whole-virus lysate using in-house ELISA methods. As China ended smallpox vaccination in 1981, we stratified the participants of the two cohorts into two age groups respectively, ≤41 (born in/after 1981) and >41 (born before 1981). Our data showed that both the median magnitude and the prevalence of binding IgG responses against Vaccinia whole-virus lysate were significantly higher among participants ages >41 in the general population cohort (Fig. 1A and 1E), which is in consistence with a previous sero-epidemiological study showing that the prevalence of anti-Vaccinia neutralizing antibodies was higher among individuals born before 1980 in China [30]. Most participants (321 out of 326) in the MSM cohort were younger than 41 (Table 1), and age-matched comparisons of the median magnitude and the prevalence of anti-Vaccinia antibody responses showed no significant difference between the MSM cohort and the male participants of the general population cohort (Fig. 1A and 1E).

**Figure 1.**
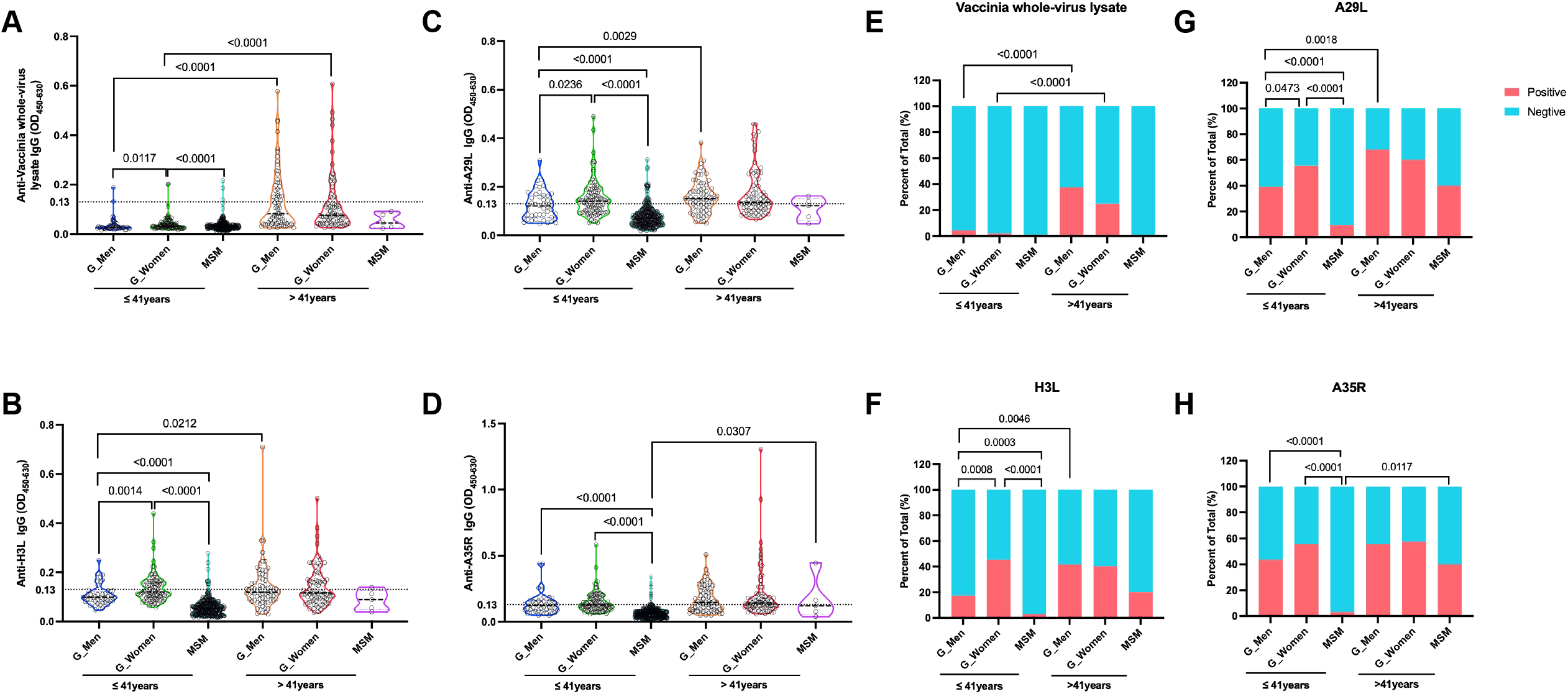
Comparisons of binding antibodies against MPXV H3L, A29L, A35R proteins and Vaccinia whole-virus lysate. The participants of both cohorts were stratified into two age groups, ≤41 years and >41 years. The participants in the general population group were further separated by sex. The median magnitude (**A** to **D**) and the positive rates (**E** to **H**) of binding IgG responses against MPXV H3L, A29L, A35R proteins and Vaccinia whole-virus lysate were compared among groups. Statistical analyses were performed by the method of non-parametric t test (**A** to **D**) or Chi-square test (**E** to **H**). G_Men (general men), n=46; G_Women (general women), n=97; MSM, n=321.

Preexisting binding IgG antibodies against MPXV H3L, A29L and A35R proteins were also detectable in both cohorts. In the general population cohort, the median magnitudes (Fig.1B, 1C and 1D) and the positive rates (Fig. 1F, 1G and 1H) tended to be higher among participants ages >41. Significant differences of binding antibody responses against H3L and A29L between the two age groups were observed for the male participants (Fig. 1B, 1C, 1F and 1G). Age-matched comparisons revealed that the median magnitudes (Fig.1B, 1C and 1D) and the positive rates (Fig. 1F, 1G and 1H) of antibody responses against MPXV H3L, A29L and A35R proteins were significantly higher among the participants ages ≤41 of the general population group than those among the participants ages ≤41 of the MSM cohort.

### Significantly higher levels of neutralizing antibody responses against Vaccinia Tiantan virus was observed in the MSM cohort than in the general population cohort

We measured the neutralizing antibody responses against Vaccinia Tiantan virus using a method of plaque reduction neutralizing test. Previous studies suggested that smallpox vaccine could confer appreciable protection against MPXV [31, 32]. Therefore, we speculate that the neutralizing antibody responses against Vaccinia Tiantan virus may reflect the potential neutralizing activities against MPXV. Our results showed that the virus inhibition ratios (%) of sera measured at a dilution of 1:30 were statistically higher among participants ages >41 than those among participants ages ≤41 in the general population cohort (Fig. 2A). Meanwhile, the percentages of individuals with a PRNT_50_ ≥30 were also significantly higher among participants ages >41 than those among participants ages ≤41 in the general population cohort (Fig. 2B). Quite unexpectedly, we found that individuals ages ≤41 in the MSM cohort showed significantly higher levels of neutralizing antibody responses against Vaccinia Tiantan virus than individuals of the same age group in the general population cohort (Fig. 2 and Supplementary Fig.1).

**Figure 2.**
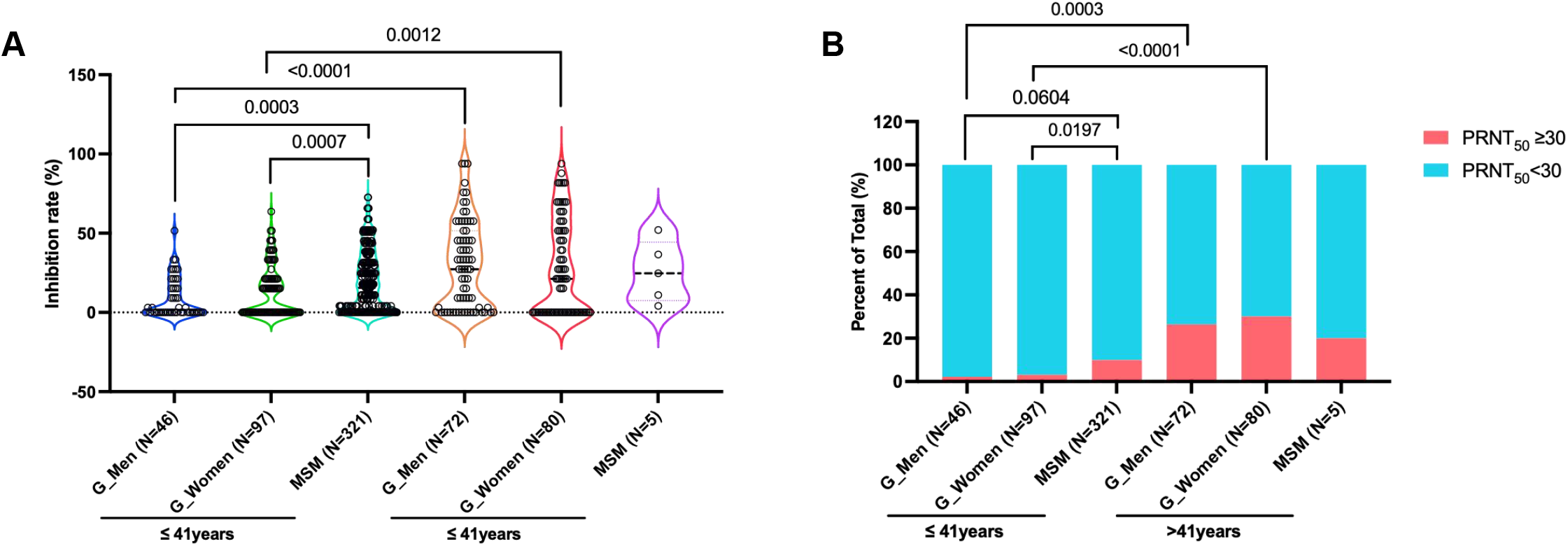
Comparisons of neutralizing antibody responses against Vaccinia Tiantan virus. The neutralizing antibody responses against Vaccinia Tiantan virus were tested using a method of plaque reduction neutralizing test. The inhibition ratios (**A**) and the percentages of individuals with a PRNT_50_ ≥30 (**B**) were compared among groups. Statistical analyses were performed by the method of non-parametric t test (**A**) or Chi-square test (**B**).

### The binding antibody responses against MPXV H3L, A29L and A35R were not associated with the binding and neutralizing antibody responses against Vaccinia Tiantan virus among participants ages ≤41 in both cohorts

To investigate whether the anti-MPXV antibody responses detected in the two cohorts were due to previous exposures to Orthopoxviruses, we analyzed the positive and negative coincidence rates between the anti-MPXV protein antibody responses and the anti-Vaccinia antibody responses. Our data showed that the positive rates of binding antibody responses against MPXV H3L, A29L, A35R and Vaccinia were significantly higher among participants with a PRNT_50_ ≥30 in the age >41 group of the general cohort (Fig.3A). In addition, the positive rate of binding antibody response against MPXV A35R was also significantly higher among individuals with a positive anti-Vaccinia binding antibody response in this group (Fig. 3B). These observations were in consistence with the findings of a mouse experiment, which showed that Vaccinia induced cross-reactive antibody responses against MPXV H3L, A29L and A35R correlated significantly with the binding antibody responses against Vaccinia whole-virus lysate (Supplementary Fig.2). These data collectively suggested that the anti-MPXV and anti-Vaccinia antibodies observed among the individuals ages >41 might partially attribute to smallpox vaccination. In contrast, no significant association between the anti-MPXV binding antibody responses and the anti-Vaccinia antibody responses was observed among individuals ages ≤41 of both the general population (Fig. 3C and 3D) and the MSM (Fig. 3E and 3F) cohorts, which implied that the preexisting antibody responses against MPXV and Vaccinia observed among individuals ages ≤41 were unlikely induced by exposures to Orthopoxviruses.

**Figure 3.**
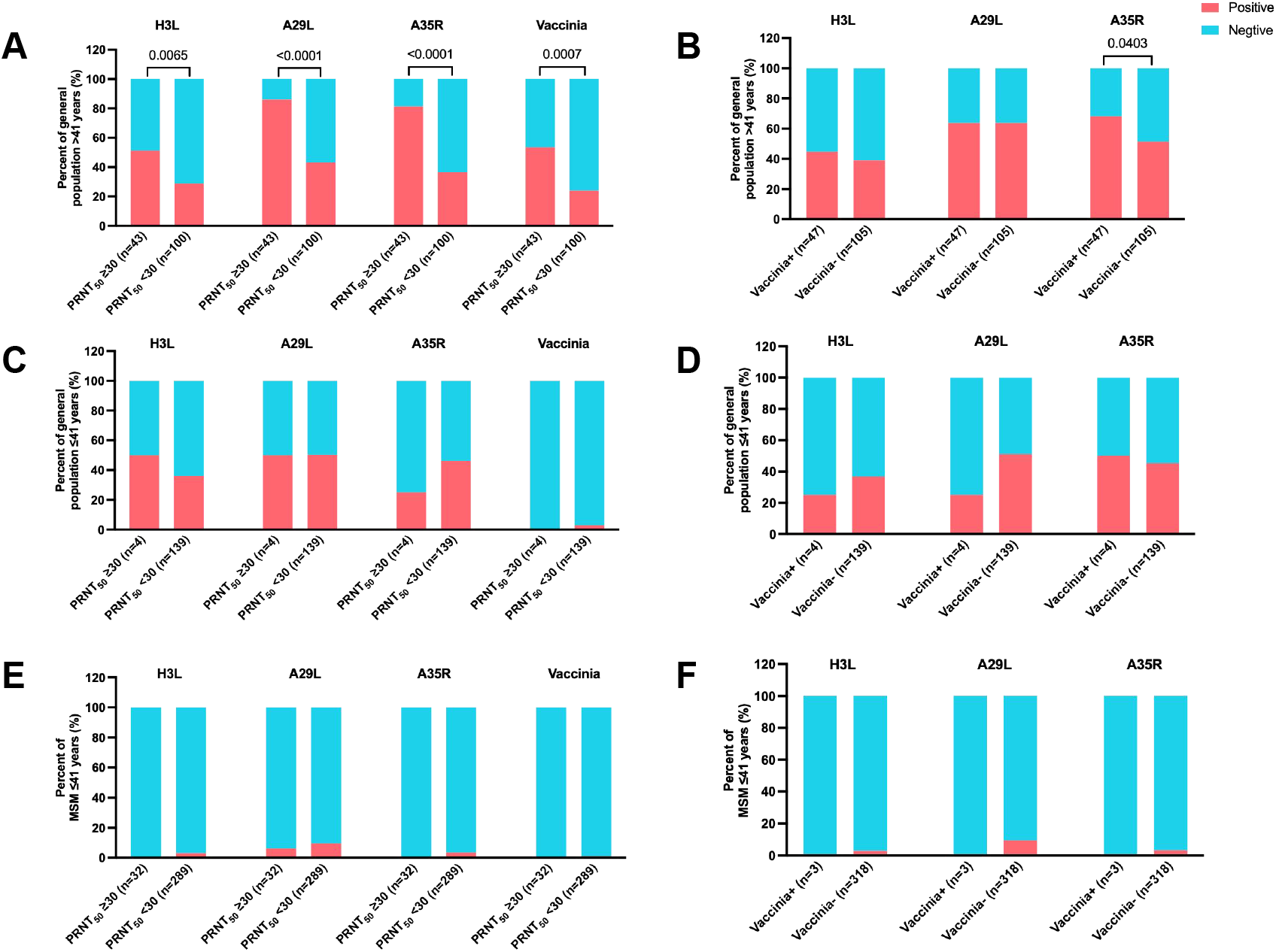
The binding antibody responses against MPXV H3L, A29L and A35R were not associated with the binding and neutralizing antibody responses against Vaccinia Tiantan virus among participants ages ≤41. The positive and negative coincidence rates were analyzed by method of Chi-square test. (**A, C** and **E)** Coincidence analyses between anti-MPXV/Vaccinia binding antibody responses and anti-Vaccinia neutralizing antibody responses in different groups. **(B, D** and **F)** Coincidence analyses between anti-MPXV binding antibody responses and anti-Vaccinia binding antibody responses in different groups.

Furthermore, to find out the potential cause of the higher anti-Vaccinia neutralizing responses observed among the participants ages ≤41 in the MSM cohort, we analyzed the associations between the preexisting anti-Orthopoxvirus antibody responses and the diagnosed sex transmitted infections (STIs) in the MSM cohort. Our data showed that the positive rates of both the binding and the neutralizing antibody responses were comparable between individuals with and without diagnosed STI infections during 6 months prior to the enrollment (Table 2).

**Table 2.** Association analyses of sex transmitted infections and anti-Orthopoxvirus antibody responses among gay men ages ≤41 y

|  | Without STI (n=275) |  | With STI (n=20) |  | P value |
| --- | --- | --- | --- | --- | --- |
|  | Negative (n) | Positive (n) | Negative (n) | Positive (n) |  |
| **H3L binding IgG** | 267 | 8 | 19 | 1 | 0.4732 |
| **A35R binding IgG** | 266 | 9 | 19 | 1 | 0.5100 |
| **A29L binding IgG** | 250 | 25 | 17 | 3 | 0.2921 |
| **Vaccinia binding IgG** | 274 | 1 | 19 | 1 | 0.1312 |
| **Neutralizing antibody** | 246 | 29 | 18 | 2 | 0.6480 |

**Note:** With STI, participants with diagnosed Syphilis, Gonorrhea, Chlamydia, genital herpes or genital warts during the 6 months before the enrollment. Without STI, participants without diagnosed STI during the 6 months prior to the enrollment. Participants who were not sure whether they had STIs or not during the 6 months prior to the enrollment were excluded from this analysis. PRNT<sub>50</sub> $\geq 30$ is classified as a positive neutralizing antibody response.

Collectively, the above data indicated the anti-Orthopoxvirus antibody responses observed among the participants ages ≤41 might not be associated with exposures to Orthopoxviruses. And the higher levels of anti-Vaccinia neutralizing antibody responses were not associated with STIs in the MSM cohort.

## Discussion

Orthopoxviruses have a broad host spectrum, ranging from humans to domestic and wild animals [33]. Variola, Vaccinia, VACV-like Brazilian isolates, Cowpox, Buffalopox and Monkeypox have been found to be able to infect human [34]. MPXV is highly identical to Variola at the genetic level [35-37], therefore, smallpox vaccines can provide substantial cross-protection against MPXV [31, 32, 38, 39]. It is speculated that the gradually increased monkeypox virus infection over time may be related to the increase in the proportion of the population that has not been vaccinated against smallpox [7, 40, 41]. A recent real-world study confirmed that receipt of 1 or 2 doses of a smallpox vaccine significantly reduced the risk for MPXV [42], although the duration of the protection was unknown.

Most confirmed cases in the current MPXV outbreak are men who have sex with men at a median age of 41 and it has been proposed that behavioral factors might contribute to the rapid transmission in this community [16]. While, it is unknown whether and how the preexisting immunities may influence the spread of MPXV-2022. In this study, we measured the anti-MPXV and anti-Vaccinia antibody responses in a cohort of gay men and a cohort of general population. Most of the enrolled gay men were younger than 41, which resembled the demographical characteristics of the confirmed MPXV cases.

Given that China eradicated smallpox in 1979 [43] and ceased smallpox vaccination since 1981, individuals born in/after 1981 (≤41) should theoretically be immunologically naïve to MPXV and Vaccinia virus. However, our data showed that binding antibodies against MPXV H3L, A29L, A35R proteins and Vaccinia whole-virus lysate could be readily detected among participants ages ≤41, although the positive rates were lower than those among participants ages >41. These findings are in accord with previous studies demonstrating that anti-Orthopoxvirus antibodies could be detected in individuals without previous infection or vaccination [21, 23].

Moreover, our data showed that the levels of anti-Vaccinia neutralizing antibody responses among the gay men ages ≤41 were statistically higher than those among the general population ages ≤41. The anti-vaccinia neutralizing antibodies observed among participants ages ≤41 were unlikely induced by unaware exposures to Orthopoxviruses, because the anti-Vaccinia neutralizing antibody response was found to be associated with the anti-Vaccinia binding antibody response in individuals who might get vaccinated against smallpox (>41) but not in those ≤41. In addition, our data elucidated that the anti-Vaccinia neutralizing antibody responses were not associated with prior STIs, which dismissed the possibility that the higher levels of neutralizing antibody responses among the gay men ages ≤41 were induced by STIs. Further investigations are required to track the origin of these naturally occurring anti-vaccinia neutralizing antibodies.

A major limitation of this study is that neutralization of Monkeypoxvirus are not detected, because the virus is not available to us. Nonetheless, our study showed that anti-MPXV and anti-Vaccinia antibodies could be readily detected in an MSM cohort and a general population cohort. And a higher level of anti-Vaccinia neutralizing antibody responses was observed among individuals who did not get vaccinated against smallpox in the MSM cohort compared to age-matched individuals in the general population cohort. Our findings highlight that the basic reproduction number (R_0_) of MPXV might be underestimated by previous studies [44, 45], because the effect of the naturally occurring antibody responses is not taken into consideration.

## Data Availability

All data produced in the present work are contained in the manuscript

## Author contributions

YW, HT and WZ designed and supervised the study; YF, HT and SL coordinated the enrollment of study cohorts and sample collection; YZ, YW and CG conducted the antibody detections; YW, YZ, YF and WW performed the data analysis; YW and YZ drafted the manuscript; WZ and YW revised the manuscript.

## Acknowledgments

We thank the volunteers who participated in this study. This work was supported in part by the National Natural Science Foundation of China (Grant No. 32270986, 81971559) and the Science and Technology Commission of Shanghai Municipality (21NL2600100).

## Figure legends

**Supplementary Figure 1.**
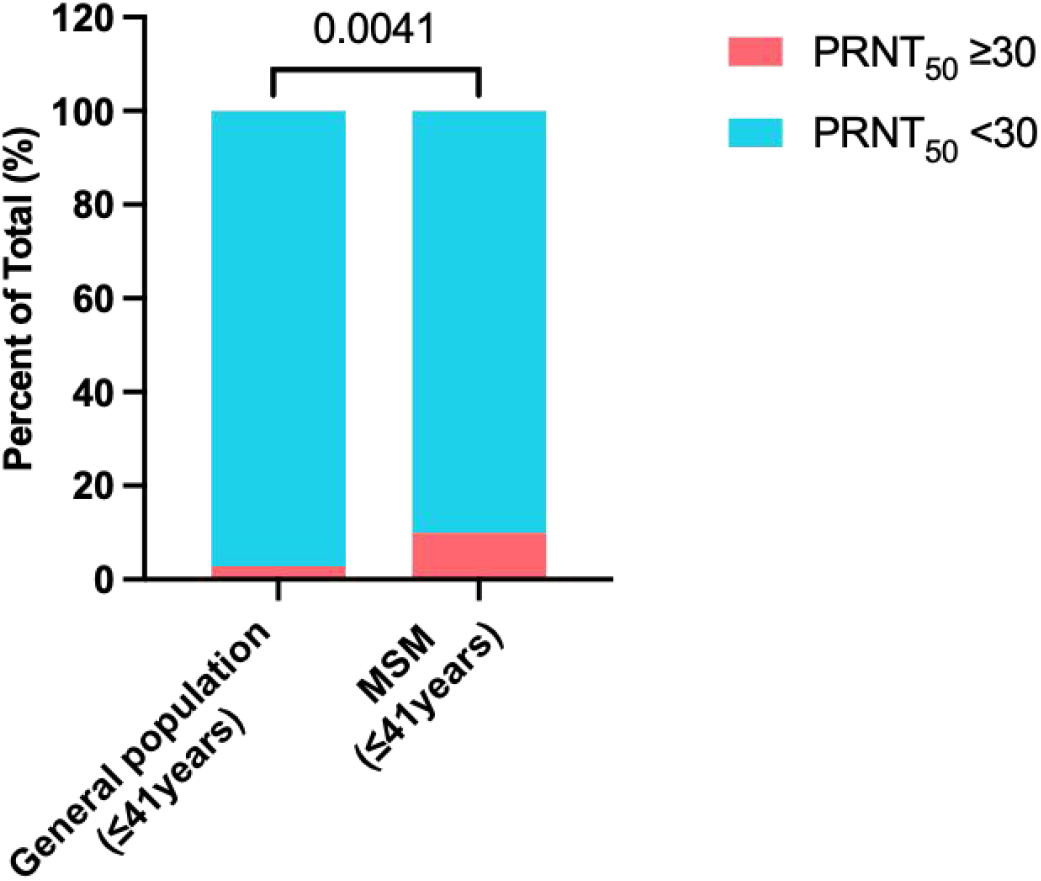
Comparison of anti-Vaccinia neutralizing antibody responses in participants ages ≤ 41 between the MSM and the general population cohorts. The percentages of individuals with a PRNT_50_ ≥30 were compared between the gay men ages ≤41 and the general individuals ages ≤41. Statistical analysis was performed by the method of Chi-square test.

**Supplementary Figure 2.**
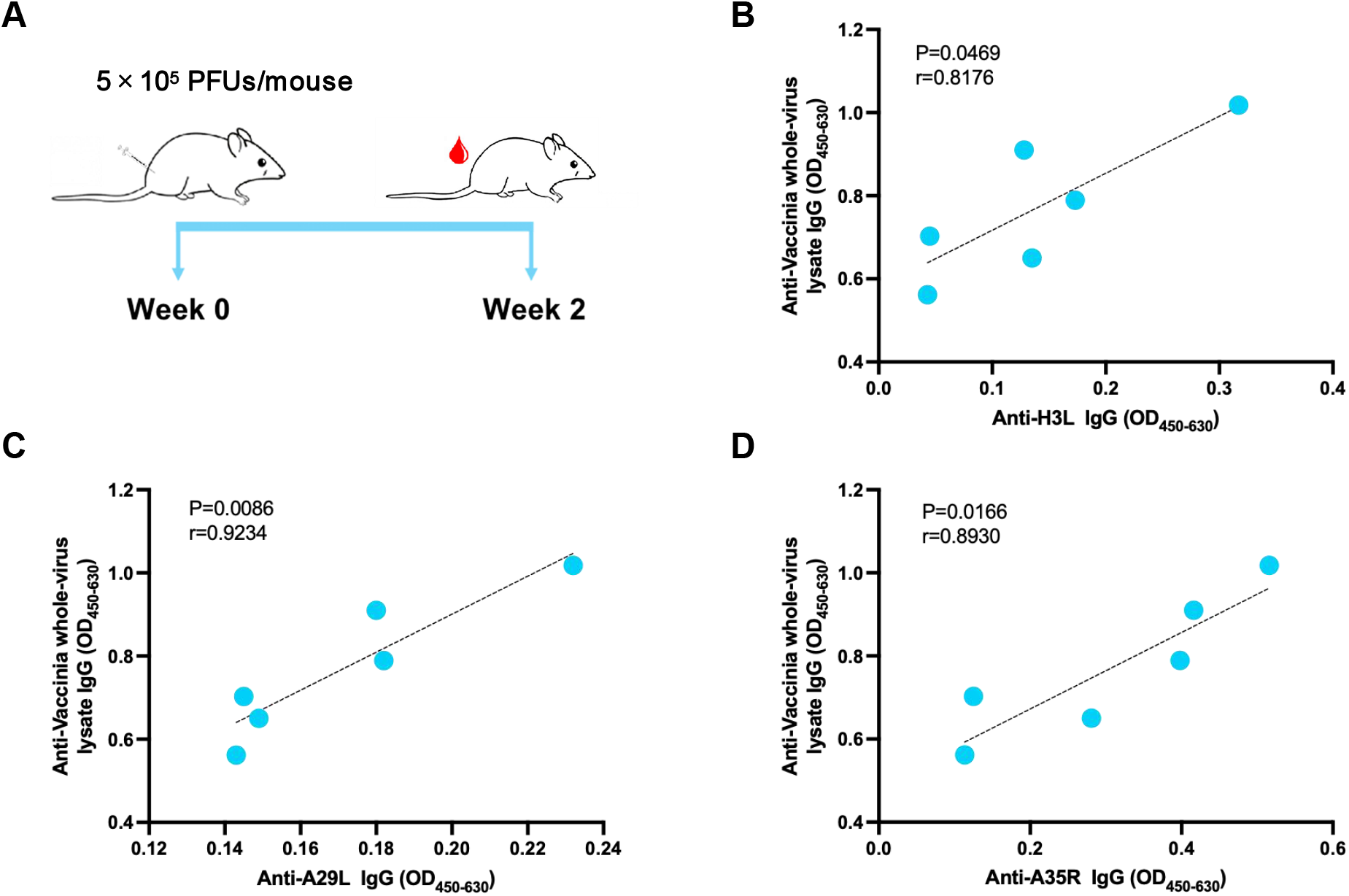
Correlation analyses of the anti-MPXV binding antibody responses and the anti-Vaccinia binding antibody responses in mice immunized with Vaccinia Tiantan virus. Female BALB/c mice were immunized intramuscularly with Vaccinia Tiantan virus (5×10^5^ PFUs/mouse) and peripheral blood was collected at 2 weeks post vaccination (**A**). Correlations between Vaccinia Tiantan virus induced cross-reactive antibody responses against MPXV H3L, A29L, A35R and antibody responses against Vaccinia whole-virus lysate were analyzed by the method of Person correlation (**B** to **D**).

